# Older Adults and Healthcare Professionals’ Perspectives on Comfort, Dignity, and Safety During Technology-Assisted Transfers: A Qualitative Descriptive Study

**DOI:** 10.64898/2026.11.17.26368333

**Authors:** Nathalie Todam Nguepnang, Sathya Karunananthan, Karine Toupin-April, Martin Bilodeau, Jeffrey Jutai

## Abstract

**Purpose:** The experience of comfort, dignity, and safety during the use of transfer technologies remains poorly understood despite evidence supporting their effectiveness in reducing work-related musculoskeletal injuries among healthcare professionals. This study explored how older adults and healthcare professionals understand and experience comfort, dignity, and safety during technology-assisted transfers.

**Materials and Methods:** A qualitative descriptive study guided by reflexive thematic analysis was conducted across three healthcare facilities in Ottawa, Canada. Semi-structured interviews were conducted with 15 older adults and long-term care residents requiring technology-assisted transfers, and four focus group discussions were conducted with 15 healthcare professionals. Data were analyzed using Braun and Clarke’s reflexive thematic analysis.

**Results:** Findings revealed an overarching theme: the relational nature of the transfer experience. Comfort, dignity, and safety were experienced as interconnected and reciprocal rather than as distinct outcomes. Three dimensions were identified: (1) comfort, which was physical, emotional, and relational; (2) dignity, which was restored through how staff treated the person; and (3) safety, which depended on trust in the person operating the device and, for professionals, on clinical judgment and adaptability. Participants also highlighted the importance of staff training, equipment customization, communication, and adequate staffing to enhance transfer experiences.

**Conclusion:** Comfort, dignity, and safety during technology-assisted transfers are not inherent properties of transfer devices but are co-constructed through interactions among older adults, healthcare professionals, and technologies. Transfer quality depended on relational practices, such as communication, privacy, and staff continuity, alongside technical competence. Training that combines technical and relational skills, stable staffing, and adaptable equipment may improve transfer experiences.

## 1. Introduction

According to the World Health Organization (WHO), the number of people aged 60 and over is expected to increase from one billion in 2020 to 2.1 billion by 2050. The population aged 80 and over is expected to nearly triple during this same period (WHO, 2025). The natural aging process inevitably leads to a decline in functional abilities, which can result in older adults being admitted to hospitals and long-term care facilities more frequently. In these facilities, mobility impairments become more prevalent, and the risk of falls and related injuries increases accordingly (WHO, 2021). Falls are the leading cause of injury among older adults, and assisting them with movement exposes healthcare professionals to a comparable risk of musculoskeletal injury. Prevalence estimates for low back pain exceed 40% among nurses and nursing aides in long-term and home care settings (Davis & Kotowski, 2015; Martimo et al., 2008). Therefore, the use of assistive technologies, such as lifts (floor, total, mechanical, and ceiling), to facilitate mobility and transfers (e.g., from a bed to a wheelchair or toilet, and vice versa), has become common in these settings. There is evidence supporting the effectiveness of these technologies in reducing musculoskeletal injury among healthcare professionals (Hegewald et al., 2018; Martimo et al., 2008). However, little is known about how their use is experienced in terms of comfort, dignity, and safety. The existing literature on transfer technologies primarily considers the perspectives of healthcare professionals and employers. These studies evaluate transfer technologies based on factors such as load reduction, injury rates, and cost-effectiveness (Hegewald et al., 2018; Koppelaar et al., 2009; Martimo et al., 2008). The few studies that have addressed patient-level outcomes have generally focused on transfer time and general comfort ratings. For example, ceiling lifts have been associated with shorter transfer times and higher comfort ratings than floor-based lifts (Alamgir et al., 2009a; Alamgir et al., 2009b). The deeper relational, emotional, and identity-related dimensions of the transfer experience have received comparatively little attention. A scoping review mapping how comfort, dignity, and safety are conceptualized and measured across technology-assisted literature confirmed this pattern^1^: safety was the most rigorously conceptualized construct, comfort occupied an intermediate position, and dignity was the least defined and measured construct. It was mainly addressed through qualitative or ethical lenses rather than through dedicated instruments or tools, which limits their evaluation and comparison across settings.

Comfort (Kolcaba, 1994), dignity (Nordenfelt, 2003), and safety (Larsson et al., 2023) are central dimensions of person-centred care, yet they are difficult to operationalize and measure consistently in real-world contexts such as technology-assisted transfers, because they are subjective, multidimensional, and shaped by context (Clancy et al., 2021; Šaňáková & Čáp, 2019). The patient perspective is particularly underrepresented.

This gap is especially significant for older adults with mobility impairments in hospitals or long-term care facilities. For this population, assisted transfers are frequent, and issues of vulnerability, functional dependence, and loss of autonomy are heightened (Heggestad et al., 2015; Liu et al., 2025). Qualitative research on dignity in long-term care settings suggests that older residents primarily define and experience dignity in relational terms, such as privacy, respectful communication, and preserving self-worth, rather than through clinical or technical criteria alone (Fuseini et al., 2023; Liu et al., 2025; Nordenfelt, 2003).

Little is known about how older adults and healthcare professionals jointly understand and experience these concepts during technology-assisted transfers. Informed by the Person-Centered Practice Framework (McCormack & McCance, 2006, 2017), this study aimed to explore how older adults and healthcare professionals understand and experience comfort, dignity, and safety during technology-assisted transfers.

## 2. Methods

### 2.1 Study Design

We conducted a cross-sectional qualitative descriptive study (Sandelowski, 2000) to explore how older adults and healthcare professionals understand and experience comfort, dignity, and safety during technology-assisted transfers. Data were collected through semi-structured individual interviews with patients and long-term care (LTC) residents, as well as focus group discussions (FGDs) with healthcare professionals, all conducted in person. The reporting follows the Consolidated Criteria for Reporting Qualitative Research (COREQ; Tong et al., 2007), and the completed checklist is provided in Supplementary File 1.

### 2.2 Settings, Participants, and Sampling

Using a purposive convenience sampling strategy, participants were recruited from three healthcare facilities in Ottawa, Ontario, Canada: Élisabeth Bruyère Hospital, Saint-Vincent Hospital (Bruyère), and Perley Health. These sites were selected because they provide continuing and long-term care and rely extensively on technology-assisted transfers. This provided a rich and varied context in which to examine everyday transfer practices. Perley Health, one of the largest long-term care facilities in Ontario, provided access to a wide variety of residents and transfer situations. Transfers at these sites were performed with ceiling lifts, sit-to-stand lifts, and floor lifts, within facility policies such as two-person transfer requirements.

Two groups of participants were included. The first group consisted of older adults and LTC residents with mobility impairments (n = 15) who required at least two assisted transfers per day using a transfer device. Individuals with cognitive impairment were not included, as this study was not designed to adapt its consent process and data collection methods to this population. The second group included healthcare professionals directly involved in patient transfers (n = 15). This group included physiotherapists, physiotherapy assistants, registered nurses, registered practical nurses, and personal support workers (PSW) or personal care attendants (PCA).

All individuals who met the eligibility criteria were invited to participate during the recruitment period, to ensure the inclusion of those with direct, regular experience with technology-assisted transfers. Those who expressed interest and were available were enrolled, and sampling prioritized availability and willingness rather than deliberate variation in participant characteristics. Recruitment procedures varied slightly by site. At Saint-Vincent Hospital, clinical or unit managers distributed study information to eligible staff members, who were invited to contact the research team; at Élisabeth Bruyère Hospital, the same approach was attempted for staff without success (see Section 2.3). The research team received contact information for interested patients or residents and then contacted them for further screening and potential inclusion in the study. At Perley Health, staff were approached via an institutional recruitment email, and residents were approached via a recruitment flyer posted in common areas. The flyer outlined eligibility criteria and study procedures. Interested individuals contacted NT directly who screened them for eligibility, obtained written informed consent, and scheduled interviews or focus groups.

### 2.3 Data Collection

Between January and August of 2025, NT collected data through in-person focus group discussions (FGDs) and individual semi-structured interviews with both groups of participants. All 15 patients and LTC residents across the three sites were interviewed individually. To accommodate the schedules of healthcare professionals, four focus group discussions (FGDs) were conducted: two at Saint-Vincent Hospital (one with RN/RPNs, n = 3; one with PCAs, n = 3) and two at Perley Health (one with PSWs, n = 4; one with a mixed group of PTs/PTAs and RNs/RPNs, n = 5) in conference rooms at the respective facilities. No healthcare professionals were recruited at Élisabeth Bruyère Hospital, and no FGDs were held there, because site managers were unavailable to respond to the recruitment emails needed to facilitate staff recruitment; only patients and residents were interviewed at this site. Interviews lasted between 30 and 45 minutes, and FGDs lasted between 45 and 60 minutes. Each participant took part in a single session; no repeat interviews were conducted.

The interviews and focus group discussions (FGDs) followed pre-established, pilot-tested guides that addressed the following: (1) experiences with transfer technologies, (2) perceived comfort, dignity, and safety during technology-assisted transfers, and (3) suggestions for improving the transfer experience. Separate guides were used for patients/residents and for healthcare professionals, following recommended practices for developing semi-structured interview guides (see Supplementary Files 2 and 3; Kallio et al., 2016). All sessions were audio-recorded and transcribed verbatim. Field notes were also taken to capture nonverbal cues not evident in the recordings. No one other than the participant(s) and NT was present during data collection. Data collection took place within the participating healthcare facilities, either in patients’ or residents’ rooms or in a conference room for staff FGDs.

Prior to participation, sociodemographic data was collected (age, sex, and length of stay for patients/residents; age, sex, professional role, and years of experience with technology-assisted transfers for professionals) through a written form. All transcripts and verbatim excerpts were anonymized using unique participant identifiers (e.g., P01 and S01 for patients and staff, respectively) before analysis and reporting. Transcripts were not returned to participants for comment or correction.

### 2.4 Research Team, Reflexivity, and Analytic Rigor

NT, a woman physiotherapist and PhD candidate with over six years of experience in qualitative health research, conducted all interviews and focus group discussions (FGDs). NT had no prior clinical or professional relationship with participants and was introduced as an external researcher. Participants were informed that the study is part of a broader doctoral project aiming to identify a way of evaluating the comfort, dignity, and safety of transfer technologies; the analysis reported here focuses on participants’ own understandings and experiences of these concepts. Participants were informed verbally and in writing about the study purpose, procedures, and voluntary nature of participation prior to providing consent.

Consistent with the principles of reflexive thematic analysis (Braun & Clarke, 2006, 2021), reflexivity was maintained throughout data collection and analysis. NT kept a reflexive journal and analytic memos to document evolving assumptions, methodological decisions, and interpretations. During interviews and FGDs, participants’ accounts were summarized back to them by NT to verify her understanding of their experiences and meanings. This in-session verification did not constitute formal member checking, as transcripts and interpretations were not returned to participants.

Regular analytic discussions among the research team (NT, JJ, and MB) encouraged critical engagement with the data and exploration of alternative interpretations. Differences in interpretation were viewed as analytically valuable opportunities to deepen understanding rather than discrepancies requiring consensus. Analytic rigour and transparency were further supported through maintenance of an audit trail documenting methodological decisions, refinements to interview guides, and the development of codes and themes throughout the study.

### 2.5 Data analysis

All transcripts were anonymized using unique participant codes (P01 for patients and residents and S01 for healthcare staff, for example). The analysis combined deductive and inductive approaches. Deductive coding was informed by the predefined domains and concepts embedded in the interview and focus group discussion (FGD) guides. Consistent with the study’s aim of exploring participants’ understanding of and experiences with comfort, dignity, and safety, data were analyzed using reflexive thematic analysis (Braun & Clarke, 2006).

NVivo was used for data management and for initial coding; because the coauthors did not have access to NVivo, coding was then continued in Word tables so that all team members could review it. The analysis followed Braun and Clarke’s six phases. NT led the coding, and the research team members (JJ and MB) engaged in regular analytic discussions to refine and deepen interpretation throughout.

#### 1. Familiarization with the Data

NT transcribed all recordings and immersed herself in the corpus by repeatedly reading the transcripts. Early analytic notes captured first impressions, emerging patterns, and sensitizing concepts drawn from the interview and focus group discussion (FGD) guides.

#### 2. Generating Initial Codes

NT systematically coded the entire dataset, identifying semantic and latent meanings related to the participants’ experiences with transfer technologies and their perceptions of comfort, dignity, and safety. Deductive codes, informed by the interview and focus group discussion (FGD) guides, were combined with inductive codes that emerged directly from the data.

#### 3. Constructing Initial Themes

Codes were organized into preliminary themes that reflected patterned meanings across interviews and focus group discussions (FGDs). This stage involved grouping related codes and exploring their conceptual connections to person-centered transfer experiences.

#### 4. Reviewing and Refining Themes

Themes were developed through an iterative process of reflexive interpretation, discussion among the research team (NT, JJ, and MB), and engagement with the data.

#### 5. Defining and Naming Themes

Themes and subthemes were clearly defined and named to reflect their conceptual essence.

A coding record was maintained to support transparency of theme development. Coding tables (categories, codes, descriptions, and illustrative extracts) were used as working documents to organize the data; they were shared with JJ and MB for comment and revised iteratively and were not applied as a fixed codebook or used to assess coding agreement. The final thematic structure, with illustrative codes, is provided in Supplementary File 5.

#### 6. Producing the Analytic Narrative

Themes were integrated into the analytic narrative presented in the Results section and interpreted in the Discussion section through the Person-Centered Practice Framework (McCormack & McCance, 2006, 2017), whose constructs are prerequisites, the care environment, care processes, and person-centered outcomes, and, for the dignity findings, through a relational dignity framework (Nordenfelt, 2003; Jacobson, 2009; Heggestad et al., 2015). Both frameworks were applied after the themes had been developed.

#### Sample adequacy and Information Richness

Data collection continued until the dataset was rich and diverse enough to allow meaningful thematic interpretation of participants’ experiences. By approximately the twelfth interview, the research team observed strong recurrence of patterned meanings across participant accounts. Nonetheless, consistent with the recommendation to interpret saturation cautiously in small, purposively sampled qualitative studies (Saunders et al., 2018), all interviews and FGDs were analyzed to maximize interpretive depth and representation of perspectives.

### 2.6 Ethical Considerations

This study was approved by the Bruyère Health Research Ethics Board (Approval *REB M16-24-03*) and the University of Ottawa Research Ethics Board (Approval *REB H-01-24-10068*). All participants provided written informed consent prior to data collection. Confidentiality was maintained by assigning unique participant codes to all data and storing recordings, transcripts, and consent forms in a locked cabinet at the Bruyère Research Institute, in accordance with institutional requirements.

## 3. Results

This section presents the characteristics of the participants and the thematic findings that address the objective of the study, which is to explore how older adults and healthcare professionals understand and experience comfort, dignity, and safety during technology-assisted transfers. The findings are organized around one overarching theme: the relational nature of the transfer experience. Within this theme, comfort, dignity, and safety emerge as interconnected dimensions rather than independent constructs (Section 3.2). Themes were developed from the data; their relationship to the Person-Centered Practice Framework (McCormack & McCance, 2006, 2017), which was applied after theme development, is examined in Section 4.2. Within each dimension, the patient/resident perspective is presented first, followed by the healthcare professional perspective. A short synthesis highlighting convergence, divergence, or relational tension between the two groups closes each section. Section 3.3 then presents participants’ preferences and recommendations for practice.

### 3.1 Participants characteristics

A total of 31 individuals were approached across the three sites. One individual declined participation after reviewing the study information and data collection procedures, resulting in a refusal rate of 3.2%. The final sample included 30 participants: 15 patients and LTC residents (Perley Health, n = 5; Élisabeth Bruyère Hospital, n = 3; Saint-Vincent Hospital, n = 7), and 15 healthcare professionals (Perley Health, n = 9: physical therapist (PT)/physical therapy assistant (PTA), n = 3; registered nurse (RN)/registered practical nurse (RPN), n = 2; personal support worker (PSW), n = 4; Saint-Vincent Hospital, n = 6: RN/RPN, n = 3; personal care assistant (PCA), n = 3). No healthcare professionals were recruited at Élisabeth Bruyère Hospital because site managers were unavailable to respond to the recruitment emails needed to facilitate staff recruitment; therefore, participation was limited to patients/residents.

The detailed characteristics of the participants are presented in Tables 1 and 2.

**Table 1.** Sociodemographic Characteristics of Patients and LTC Residents (n = 15)

| **Characteristic** | **Patients and LTC residents (n = 15)** |
|---|---|
| **Setting / n** |  |
| Perley Health | 5 |
| Saint-Vincent Hospital | 7 |
| Élisabeth Bruyère Hospital | 3 |
| **Sex/ n** |  |
| Female | 6 |
| Male | 9 |
| **Age group (years), n** |  |
| 51-60 | 2 |
| 61-70 | 4 |
| 71-80 | 7 |
| Over 80 | 2 |
| **Main medical condition / n** |  |
| Stroke | 2 |
| Hip or femur fracture, or hip arthroplasty | 3 |
| Bariatric care needs | 2 |
| Age-related functional decline or mobility limitation | 3 |
| Other neurological, limb-related, or chronic conditions | 5 |
| **Length of stay, median (range; n = 13 reported)** | About 12 months (2 months to 5 years) |
| **Weight, kg, range (n = 12 reported)** | 54-279 |
| **Not reported / n** | Length of stay, 2; weight, 3 |

**Table 2.** Professional Characteristics of Healthcare Staff (n = 15)

| **Characteristic** | **Healthcare professionals (n = 15)** |
|---|---|
| **Role / n** |  |
| Physical therapist (PT) or physical therapy assistant (PTA) | 3 |
| Registered nurse (RN) or registered practical nurse (RPN) | 5 |
| Personal support worker (PSW) | 4 |
| Personal care assistant (PCA) | 3 |
| **Sex / n** |  |
| Female | 12 |
| Male | 3 |
| **Age group (years) / n** |  |
| 30-40 | 3 |
| 41-50 | 6 |
| 51 or older | 4 |
| Not reported | 2 |
| **Experience with patient transfers, median (range; n = 13 reported)** | 10 years (1.5 to 27 years) |
| **Setting, n** |  |
| Perley Health | 9 |
| Saint-Vincent Hospital | 6 |

#### 3.1.1 Profile of Patients and LTC Residents

Most of the participants in this group were older adults, with the largest group being between 71 and 80 years old. They presented a broad range of mobility impairments and neurological conditions, including stroke, hip fracture, cerebral palsy, multiple amputations, spinal cord injury, multiple sclerosis, and age-related functional decline. Bariatric care needs were particularly notable in the hospital setting, affecting two individuals (Table 1).

Length of stay varied substantially, ranging from short-term rehabilitation periods of a few months to long-term residency exceeding five years. This reflects both transitional and permanent care trajectories. Weight profiles also varied widely, highlighting the heterogeneity of physical assistance needs during transfers. Overall, the patient sample captured a realistic cross-section of long-term care (LTC) and complex care populations.

#### 3.1.2 Profile of Healthcare Professionals

The healthcare professional group was composed of a mix of registered nurses (RNs/RPNs), personal support workers (PSWs), personal care assistants (PCAs), and rehabilitation clinicians (PTs and PTAs). The staff represented a wide range of ages, with most between 30 and 50 years of age (Table 2).

Their experience with patient transfers ranged from 1-3 years of recent clinical experience to more than 25 years of hands-on practice. This diversity ensured representation from frontline caregivers, who perform transfers multiple times daily, and rehabilitation professionals, who engage in specialized mobility interventions. These interventions include assigning specific transfer technologies to patients or residents upon admission to a healthcare facility depending on their level of functional independence and needs. Together, the staff provided a comprehensive view of transfer practices shaped by their varied professional backgrounds, clinical responsibilities, and levels of expertise.

### 3.2 The Relational Dimension as a Cross-Cutting Thread

A core finding of this study is that comfort, dignity, and safety are not purely technical aspects of the transfer, but rather, they are influenced by the quality of the relationship between patients and staff throughout the process. Participants in both groups consistently described a “human factor”: communication, eye contact, and verbal “check-ins” performed by staff during the procedure. These factors determine the quality of the transfer, regardless of the equipment used. When this relationship is strong, patients report feeling “in charge”, even while suspended in a mechanical device. For example, P02 said, *“The effectiveness of the staff is 100% related to how safe I feel.”* Conversely, even technically adequate equipment fails to produce a positive experience when the relational dimension is absent, as noted by P02 regarding dignity: *“I don’t think there’s anything about the equipment that lends to dignity. There is nothing. There is nothing. There is no dignity in being lifted naked in the sling! There is no dignity.”* This relational thread connects the three dimensions : comfort, dignity and safety, examined below.

#### 3.2.1 Perceptions and Definitions of Comfort : Comfort as a Relationally Mediated State

Within this relational frame, comfort encompassed physical, emotional, and personalization-related dimensions for both patients/residents and healthcare professionals.

##### The Physical Dimension of Comfort

For patients and residents, comfort is primarily defined as the absence of pain. P05 illustrates this idea: *“When there’s no pain involved […] I feel comfortable when getting moved. No pain or anything.”* Proper positioning and careful equipment handling were also identified as contributing to physical comfort. This includes adjusting slings, preventing skin pinching, and avoiding sudden movements. For example, P02 noted that discomfort during transfers was mostly due to mechanical defects, substitute equipment, or improper sizing. *“Pain free! Yes! The sling is not good for my skin. It shears my skin. That’s not good. When my sling got dirty, they brought a different one that was smaller and uncomfortable.”*

Furthermore, technical failures, such as rapidly draining lift batteries, prolonged physical suffering, as illustrated by this quote from P02: *“The battery runs out quickly. I’m in agony for five minutes instead of two”.* The same participant also noted that standardized equipment often failed to accommodate complex anatomical profiles, such as limb loss or severe obesity. This forced patients to endure ill-fitting devices. *“Nothing is made to accommodate multiple amputations.”* The idea that personalized equipment should be considered to meet individual needs instead of standardized ones is supported by P09 (bariatric care needs), who emphasized the difference between a ceiling lift with two prongs and one with four prongs, the latter of which is more suitable for bariatric individuals (see Figure S1, Supplementary File 4). *“This four-pronged lift is more comfortable than the two-pronged lift at the general hospital. It cradles you without causing pain.”*

Healthcare professionals agreed that physical comfort requires pain prevention and conceptualized this through clinical execution, ensuring precise sling placement, anatomical alignment, and pressure relief, as reported by S9 and S3, respectively.

*S9: “Not having any physical discomfort and not being in poor positioning that would make you feel awkward… Make sure that the skin is not pinched, pulled, or twisted in any way when using a device.”*

*S3: “When you’re talking about comfort, you have to position the patient properly in the chair." Sometimes, you put the patient too low, and they slide on the chair. You have to make sure the patient sits all the way back in the chair! That is comfort.”*

##### The Emotional and Psychological Dimension of Comfort

Beyond physical comfort, technology-assisted transfers can evoke feelings of profound psychological vulnerability. Patients described a grieving process associated with losing functional independence and experiencing transfers as a state of helplessness or melancholy. P01 said, *“Well, I guess I will say that I feel out of control. When you are not in control, it is a state of helplessness. That’s how I would describe it." […] It’s depressing to lose the ability to stand and walk for a long time. I call it a state of melancholy, meaning I can’t do anything on my own.”*

Patients and residents reported that emotional comfort was restored when they developed a deep sense of trust in their caregivers’ competence. For example, P15 said, *“I feel comfortable because I know what they’re doing. They’re trained for this job, so I’m comfortable with them doing it. I trust them.”*

From the perspective of healthcare professionals, emotional comfort was defined as proactively mitigating resident anxiety through step-by-step communication, seeking consent, and reading nonverbal distress indicators throughout the transfer process, as mentioned by S1 and S15. S1 said, “*They like to be talked to before being moved. It makes them feel respected and less anxious.*” S15: *“It’s pretty much communication. Like letting them know, step by step, what you’re doing. We look at their facial expressions and read their body language. So, if they make a facial expression, grunt, or move, those little cues help us determine whether we should change something.”*

##### Personalization and Relational Consistency

For both groups of participants, comfort was linked to tailoring the transfer approach to the specific needs and preferences of each patient and resident, as well as maintaining care continuity. On one hand, patients and residents feel more secure when the same person performs the transfer consistently. They believed that relief or “float” staff who were unfamiliar with their routine caused acute pain, as reported by P5: *“A couple of them don’t know how to use this thing properly, and I’m in pain when they move me! The regular PSW at night in the morning has no problem. But the one who replaces her when she’s off is painful. Just because they don’t know how to put the sling on properly.*”

On the other hand, professionals confirmed that relational familiarity is an essential prerequisite for psychological ease, as reported by S4: *“Knowing their baseline, knowing triggers, knowing a whole set of things, and having familiarity that breeds comfort and trust.”*

Overall, both groups agree that comfort requires the absence of physical pain, but they emphasize different mechanisms for achieving it: patients focus on the physical fit and technical reliability of the equipment itself (sling size, battery performance, customization for amputation or skin sensitivity), while professionals focus on the relational conditions surrounding the transfer (a calm environment, personalization, and staff familiarity). This suggests that comfort is produced by a combination of adequate equipment and relational quality, rather than by either factor alone.

#### 3.2.2 Perceptions and Definitions of Dignity

The clearest illustration of the relational thread introduced above concerns dignity. Across patient and resident accounts, transfer technology is consistently described as having no inherent dignity. Dignity during a technology-assisted transfer is not guaranteed by the technology (device) itself. Instead, it must be actively restored moment by moment through the attitude, empathy, and communication of the staff performing the transfer. This tension between mechanical execution and interpersonal care is present in the three sub-dimensions examined below: Personhood and respect, environmental privacy and modesty, and the technological boundary between human and the technology.

##### Personhood and Respectful Care

Patients and residents associated dignity with being recognized as whole human beings rather than being treated like objects or clinical tasks. They experienced violations of dignity when staff displayed impatience, handled them roughly, or dismissed them verbally, as illustrated by the following quotes:

P05: *“I keep telling her I’m not a dog! I’m a human being! Treat me like one.*”

P15 *:“ By dignity you just like, kind and careful… You don’t make a big deal out of anything.*” Similarly, healthcare professionals defined dignity as preserving patients and residents’ identities and self-worth.

S9: *“Preserving their sense of self. That would be my best stand… making sure that they feel respected as a person.*”

##### Privacy, Modesty, and Environmental Control

Conversely, the exposure of private body parts during mechanical transfers was reported by residents as the most severe threat to dignity. For example, suspension in a lift during commode or shower transfers exposed extreme bodily vulnerability: P02 *“There is no dignity in being lifted naked in the sling! Yeah, there is no dignity. If you’re naked, going to the toilet, you’re in a sling… everything is always just out there, exposed to the world.*” This was feeling supported by another resident P04 who said: *“Well, when they move me down the hall with no clothes on and find that rather awkward.*”

Professionals viewed environmental control, closing room doors, drawing privacy curtains, covering residents with blankets, and respecting cultural or religious modesty preferences as the baseline of dignified care, as reported by S3 and S15 during a FGD : *“Privacy is number one. You don’t leave the door open… you close the door. You close the curtain for the dignity of the patient*”. *They also emphasized the importance of respecting patients’/residents’ cultural sensitivities, e.g.,* providing female-only care providers for Muslim female residents, *as reported by S8 : “ Culturally, you have to be respectful… we had one lady on our unit that are Muslim, right? So it’s a religion thing. They don’t want men to take care of them… So culture thing.*”

##### The Technological Boundary: Machine vs. Human Relationship

Across the interviews and focus groups, patients and residents consistently emphasized that transfer technologies do not inherently confer dignity. They described being lifted and suspended in midair as an unavoidably mechanical and dehumanizing act, regardless of the device used. As noted by P05: *“There is nothing about the equipment that lends dignity”.* However, patients also stressed that a staff member’s attitude has the power to transform an otherwise undignified procedure. P02 captured this contrast succinctly: *“There is no dignity in being moved via a lift. However, I feel dignified by the attitude of the staff”.* This theme was particularly salient among participants who were completely dependent on others. For them, dignity was not inherent in the technology, but rather, it was conferred through interpersonal care. As P06 explained: *“When you’re incapable of doing anything yourself and someone has to do it for you there? Kind, considerate, and they understand what you’re going through, that is feeling treated with dignity.”* Participants also described specific behaviors that help preserve dignity during transfers, such as making eye contact, being patient, and checking in moment to moment. As P06 put it, “*Patience. How are you treating the person? Are you maintaining eye contact? Are you stopping to check in?”*

Healthcare professionals echoed this sentiment, noting that maintaining eye contact and providing continuous verbal guidance are essential to counteracting the unsettling experience of mechanical suspension. As one nurse, S1, explained: *“We always tell them, “Look at our eyes. Look at our eyes.”* We maintain eye contact and read their nonverbal cues and comfort level”. S9 further emphasized that, *“since being hoisted leaves residents “floating in human’s hand”, preserving dignity requires an active, relational presence; consent; and keeping the resident covered to preserve their “sense of self” rather than processing them as a mechanical task.”* Taken together, these cross-stakeholder accounts reveal a central conceptual distinction: Transfer equipment introduces a mechanical and potentially dehumanizing baseline, but dignity is restored or withheld through human interaction. The technology establishes the physical conditions of the transfer, but the staff’s relational conduct determines whether the experience is dignified.

#### 3.2.3 Safety: Physical Protection, Clinical Judgment, and Relational Trust

Safety emerged as the third nested dimension of the overarching relational thread and was universally recognized as the non-negotiable baseline of technology-assisted transfers. While institutional policies usually define safety in terms of systems-based criteria, such as equipment load limits, maintenance routines, and fall-prevention protocols, participant accounts revealed that safety is experienced and enacted relationally. For patients and residents, feeling safe depended less on the mechanical reliability of the device and more on interpersonal trust and confidence in the operator’s technique. For healthcare professionals, safety required dynamic clinical judgment, continual adaptation to residents’ changing conditions, and navigation of structural constraints, such as staffing shortages. Together, these perspectives highlight a central conceptual distinction: Systems-based safety establishes the technical conditions of a transfer, but relational safety determines how safe the transfer feels and unfolds in practice.

##### Physical Protection and Fall Prevention

Patients consistently defined safety in experiential terms, primarily as an absence of falling, pain, or collision during transfer. As one resident, P12 explained: *“Falling! You can feel unsafe if you think you are going to fall”*. Another, P02 emphasized the importance of staff competence: *“For me personally, safety is 100% related to how safe I feel, in that I won’t sustain further injuries because the staff doesn’t know what they’re doing.”*

In contrast, healthcare professionals articulated safety through a clinical and procedural lens, emphasizing correct equipment selection, routine maintenance, and proper body mechanics. As one staff member S15, summarized: *“Safety equals the right equipment for the right patient for the right reasons, causing no harm”.* These accounts illustrate how systems-based safety provides the structural foundation for preventing harm while patients rely on relational cues to assess their protection.

##### Clinical Judgment, Adaptability and Adaptability

Healthcare professionals emphasized that physical safety is not a static baseline but a dynamic clinical process that requires ongoing evaluation and real-time adaptability. Instead of treating a resident’s ability to transfer as a fixed attribute, staff explained that medical status, physical weakness, and diurnal fatigue necessitate continuous clinical adjustments. As one nurse explained: S9: “*If they’re having medical issues where they’re weaker, we would accommodate whatever transfer is necessary. You have to use your clinical judgment.”*

One PT noted that physical fluctuations often follow daily patterns or medication cycles, especially in patients with neurodegenerative conditions like Parkinson’s disease. S5 : “T*echnically, the way it’s meant to work is there is supposed to be one transfer status, but knowing that we’re all human, we have ups and downs. I often use the example of Parkinson’s where medication can be extremely effective in certain windows of times, and then you come back an hour later… it seems like a totally different person…”*.

To safely accommodate these shifts, a resident’s transfer protocol may evolve throughout the day. For example, it may transition from manual side-by-side assistance in the morning to a sit-to-stand lift in the afternoon and ultimately to a total ceiling lift in the evening when exhaustion sets in. As one nurse, S9, explained: *“If they’re having medical issues where they’re weaker, we would accommodate whatever transfer is necessary. You have to use your clinical judgment”*.

To maintain safety across shifts and communicate dynamic adjustments seamlessly to incoming care teams, staff relied on visual signage, door logos, and pictograms posted outside residents’ rooms. Together, these adaptive practices demonstrate how clinical judgment and visual signage bridge the gap between rigid institutional safety protocols and residents’ fluctuating, real-time needs.

##### Systemic Constraints and Staffing Shortages

Both patients, residents and professionals identified staffing shortages as the primary structural barrier to safety and dignity during transfers. Policies requiring two-person assistance for mechanical lifts (“No-Lift Policy”) often resulted in delays when staff were unavailable. As one *professional, S13 explained, “When you want to do a transfer, we need two people… We wait! If the patient is wet and needs to be changed, they have to wait. The equipment is good, but we need more staff”*. These accounts underscore how system-level constraints directly shape the relational experience of safety. They reveal that even well-designed equipment cannot compensate for insufficient staffing.

Overall, although both groups prioritize safety, professionals tend to focus on the prevention of harm through clinical risk-adjustment protocols, such as teamwork ratios and situational escalation, whereas patients tend to prioritize trust in a specific, known operator. Similarly to the tension identified for dignity, professionals describe safety as a system-level property (technique, staffing ratio, protocol), while patients experience it as a relational property (familiarity and competence of the individual operator).

### 3.3 Participant Preferences and Recommendations

Participants from both groups went beyond articulating the conceptual and ethical dimensions of technology-assisted transfers to offer pragmatic, experience-based recommendations for improving clinical practice and care delivery. These insights bridge the gap between mechanical execution and person-centered care. They reflect a shared desire to ensure that transfer technologies support, rather than undermine, patient’s and resident’s comfort, dignity and safety. Rather than viewing device design in isolation, patients, residents and healthcare professionals emphasized that effective technology integration requires alignment across operator competence, equipment functionality, communication workflows, and organizational support. The following sections examine the participants’ recommendations across four key domains: training and operator expertise; equipment selection and technical design; communication and coordination protocols; and staffing and relational care continuity.

#### 3.3.1 Training and Expertise

Both residents and healthcare professionals emphasized a foundational recommendation: reforming educational practices to ensure that operator competence aligns with experiential and situational care needs. Patients often cite a lack of specialized training as a primary source of pain and anxiety. One common recommendation is to implement standardized training to ensure that all staff, especially “floats” and temporary workers, handle equipment with the same level of competence as regular staff.

*P06 “ We need standardized training for them. Why do I say that so quickly? Because I’ve seen it at the rehab centre […] Standardized training for the staff is lacking here.*”

*P05 “Couple of them don’t know how to use this thing properly, and I’m in pain when they move me! […] The one that replaces her [regular PSW] when she’s off, it’s painful […] Just because they don’t know how to put the sling on properly.*”

One resident specifically suggests that staff should be required to get into a sling themselves during training to understand the physical and emotional sensation of being lifted. P03 : “ *I also think that one of the things that should happen is that the people operating this sling should be made to get into it and feel the differences.*”

Healthcare professionals emphasize this recommendation by pointing out that, while general training is available, it is often not “just-in-time.” They recommend a training-on-demand structure, in which staff receive a refresher on a specific device, such as a sit-to-stand lift, when a new resident requiring that technology arrives on the unit. *S5: “[…] I think perhaps a better structure for better availability to training on demand for specifically what is needed in the moment is probably the best. […] we get plenty of training, but we don’t always get the training that we need in the moment that we needed.*”

They also advocate for “refresher courses” to prevent the development of “bad practices” over time. S15 *“Refresher courses are very good because some of the nurses who have been here for so many years, they’re using their old ways of doing things. And then we get rusty, sometimes we keep on doing things that on old ways and yeah, so refresher courses for sure*”.

S5 “*Example: if my colleague works like in the unit where they haven’t used the Sit-to-stand lift in three months and all of a sudden there’s a new resident who requires that the sit-to stand lift, the colleague doesn’t need to be trained on all the different transfer techniques! He needs to be refreshed on the sit-to-stand lift at that time.*”

Both groups converge on the need for more frequent training, but their focus differs: patients seek standardization to ensure safety regardless of the operator, while professionals seek specificity and timing to ensure clinical competence for the diverse needs of their current patients and residents. While experiential and on-demand training enhance operator competence and establishes the human skill set necessary for safe transfers, participants emphasized that staff expertise must be supported by appropriately designed, ergonomically sound equipment.

#### 3.3.2 Equipment Choice and Technical Design

To support the staff’s expertise, patients, residents, and healthcare professionals expressed their technical preferences and suggested hardware refinements. These refinements aimed to optimize mechanical performance, minimize physical discomfort, and reduce operator strain.

Patients and residents nearly universally prefer ceiling lifts to floor-based mechanical lifts. Patients and residents prefer ceiling lifts because they allow for more precise positioning. Standalone lifts, on the other hand, can make patients and residents feel “scrunched in a ball” or “hovering” awkwardly over a chair.

*P02:* “*I would however say ceiling lifts would be 100% better than… mechanical standalone lifts. For me, a ceiling lift is almost critical because it can position me directly above where I need to be and then just drop me down.*”

*P12:* “ *I really do dislike it [floor lift]. I mean I find that your scrunch stuff in a ball too much. And it’s just uncomfortable.*”

Additionally, patients with sensitive skin or amputations recommend customized or “comfy” slings to prevent shearing and pressure sores. “

*P02:* “*I think there’s nothing that um is made specifically to accommodate multiple amputation. […] the sling is… Not good for my skin. And it shears my skin. […] In general, I’d like there to be more options of slings. Because not everyone fits the mold… I’m in agony. Every day I transfer… injuries caused by my sling*”.

Healthcare professionals also strongly recommend expanding the availability of ceiling lifts, noting that they are easier to maneuver and reduce physical stress on caregivers. They propose technical upgrades, such as integrating weight scales directly into ceiling lifts, to avoid the danger of manually attaching heavy external scales (currently only common on floor lifts).

*S14* asked “*Can the weighting system be incorporated where you can like, let’s say when you lift the patient, you can see the weight of the patient also? […] Having that weight on top of the machine [external scale], it’s something like really heavy. I’m scared that when I’m using it… it might fall on the patient.*”

This consensus on ceiling lift technology aligns with previous research on device evaluation showing that ceiling lifts are associated with shorter transfer times, higher patient comfort ratings, and lower implementation barriers when integrated early into care routines compared to floor lifts (Alamgir et al., 2009a, 2009b; Koppelaar et al., 2009).

Overall, there is strong alignment among stakeholders regarding the superiority of ceiling-mounted technology for patient comfort and staff ergonomics.

However, an optimized design and technical functionality of the equipment are insufficient if the transfer procedure lacks active and respectful communication between the operator and the resident.

#### 3.3.3 Communication and Engagement Protocols

Acknowledging that mechanical transfers have the potential to compromise a resident’s sense of autonomy, participants underlined the critical role of structured verbal and visual communication protocols in preserving that autonomy and maintaining consistency across shifts. Patients and residents recommended a “checklist” approach to communication, where staff would explain every technical step (e.g., which loop of the sling is being attached). They value being treated as partners in the transfer process.

P06: “*At the rehab centre… whatever they’re doing and they’re moving you, whether it’s OT or it’s PT, it does not matter… they’re always giving feedback on what they’re doing. So if they’re about to put the sling, they’re gonna tell you: OK, we’re going to start here. We’re going to do X, we’re doing Y, we’re going to do Z… It gives you a checklist of what you’ve done or haven’t done*”. One patient noted that if they sense the staff is inexperienced, they will "direct them" to ensure their own safety. P01: “*My priority is I hope the person who is doing it has done it before. When you sense that this person is doing it for the first time, that is when you feel a little bit apprehensive. But what I do, is I direct them. I say: ‘you do this; you do that… strap.’ If they have not done it before, then I direct them, I am in charge*”.

Healthcare professionals recommended standardizing visual aids and door signage (logos) to communicate real-time transfer protocols across shift changes. They emphasized that visual signage is an essential cognitive aid for incoming staff, allowing them to anticipate fluctuating patient needs before initiating a transfer. *S5:* “*Having those multiple logos on a wall… alerts the staff who are coming into the room that this person might not be as straightforward… so they can have a better read from the get-go in determining how they set up for a given transfer*”.

They also acknowledged using pictograms or “logos” on room doors to communicate a patient’s current transfer status (e.g., “Two-person pivot”) across shifts. S8 : “*Sometimes you might have the logos on the wall. Day shift: two staff pivot-disk transfer. Evening shift: two staff - Sara Lift… that way it at least alerts the staff who are coming into the room that this person might not be as straightforward… and that there will be these fluctuations so they can have a better read from the get-go.”* They emphasize that continuous verbal check-ins are vital for assessing a patient’s “readiness” and recognizing non-verbal indicators of distress. S8: “*Pretty much communication. Like letting know step by step what you’re doing… We look at their facial expression, we read their body language… Talking through the process, making sure they feel comfortable with you as a staff. Asking permission*”.

Both groups prioritize communication. Patients and residents view it as a way to regain control over a “helpless” situation, while professionals use it as a clinical monitoring tool to ensure safety during shift changes.

However, even the most effective communication strategies and technical skills cannot be maintained without organizational structures that support staff familiarity and relational continuity.

#### 3.4.4 Staffing and Relational Consistency

The final domain of recommendations addresses the systemic and organizational conditions required to foster relational continuity and promote dignified transfer practices. These conditions include staffing ratios and care assignment stability.

Patients and residents emphasize that consistency in staffing is a priority need. They report that safety and dignity are best maintained by regular staff who “know the file” and the patient’s specific pain points. P06: “*It’s um, you gotta have a nurse knows the file, OK or PCA or wherever. If they don’t know you from Jacob, they’re going to make mistakes. They’re going to assume that you can walk. They’re going to assume that you can support yourself, right? So what factors are important to me is that they know what they’re doing with me well before they come in*”.

*P02 :* “*When you have your regular staff, there’s no issues, they know how to handle you… when you get staff who are floats or who are temporaries, they come in and then everything’s off the table. And then it’s an awful experience because they don’t know what they’re doing*”.

Healthcare professionals explicitly state that staffing shortages are the primary barrier to a good transfer experience. They recommend increasing staff levels so residents do not have to wait in an uncomfortable or unsafe situation because a required two-person transfer cannot be performed immediately. S12 said : “A*t the same time, if I have any recommendation to say: we need more staff. What happens is because when you want to do a transfer, we need two people, your partner. And in the evening you have 11 patients, the other one has 11 patients, we have 22 for two. If you have to transfer this patient by two… you won’t finish! […] We wait! That means, if the patient is wet, and needs to be changed, the patient has to wait. The equipment is good, but we need more staff.”*

Overall, there is a relational tension where the “No lift policy,” which requires two people, is technically sound but is hindered by staffing levels in practice. This leads to a compromise in the dignity and comfort that the policy was designed to protect. Taken together, participants’ recommendations converged on four conditions for comfortable, dignified, and safe transfers: situation-specific training, adaptable equipment, structured communication, and stable staffing.

## 4. Discussion

### 4.1 Revisiting the Conceptualization of Comfort, Dignity, and Safety

The principal finding was that comfort, dignity, and safety were experienced as fundamentally relational rather than solely technological outcomes. Participants consistently described these dimensions as emerging through interactions among residents, healthcare professionals, and transfer technologies. These findings echo the call in the dignity literature to move beyond single-attribute definitions of dignity towards multidimensional, relational accounts (Clancy et al., 2021; Jacobson, 2009).

Comfort is redefined as a holistic state that involves an absence of physical pain and a presence of emotional ease. Patients and residents often express this as being “pain-free,” while professionals describe it as a state of “reassurance.” Dignity is conceptualized as preserving personhood through privacy and respect. It is maintained through small actions, such as covering the patient or knocking before entering. Consistent with qualitative findings, residents’ dignity in long-term care depends on everyday relational gestures rather than formal policy (Fuseini et al., 2023; Liu et al., 2025). Safety is clinically defined as “causing no harm” and experientially defined as the absence of falling, or any event that could cause injury. Taken together, the findings suggest the following preliminary working definition of these concepts as a triad of patient-centered security in the context of technology-assisted transfers: a state in which physical harm is prevented (safety), physical and psychological distress are absent (comfort), and the individual’s sense of self and privacy remains intact (dignity).

### 4.2 The Primacy of the Relational Dimension: A Person-Centered, Dignity-Informed Perspective

Participants rarely attributed dignity to the technology itself. Instead, dignity arose through the way technology was introduced, explained, and used by healthcare professionals. Patients and residents explicitly stated that their comfort and safety depend on staff conduct rather than the mechanical device itself.

The technology is repeatedly described as merely a “machine” or a “tool,” while eye contact, verbal check-ins, and patience redeem the inherent vulnerability of being “hoisted in the air.” The Person-Centered Practice Framework (McCormack & McCance, 2006, 2017) is the primary interpretive lens applied to the findings, situating this relational dimension within a coherent structure. Staff competence and familiarity are repeatedly identified across the comfort and safety themes (Sections 3.2.1 and 3.2.3) and correspond to the framework’s prerequisites. Environmental features, such as closing doors and curtains during transfers and maintaining adequate staffing levels (Sections 3.2.2 and 3.3.4), correspond to the care environment. Communicative practices, such as verbal check-ins, explanations, and shared pacing of the transfer, correspond to care processes (Sections 3.2.1-3.2.3). Comfort, dignity, and safety occupy the position of person-centered outcomes. They are not properties of technology but rather products of the interaction between prerequisites, environment, and process.

Considering the salience of dignity-related tensions in the dataset, particularly between technology and interpersonal care, findings relating specifically to the dignity theme require further interpretation through a relational dignity framework (Nordenfelt, 2003; Jacobson, 2009; Heggestad et al., 2015). This framework addresses how dignity is threatened or restored through the care relationship, rather than residing solely within the individual or the device (i.e. the technology). The data illustrate this distinction starkly (Section 3.2.2): a technically perfect ceiling lift transfer can result in a total loss of dignity if the operator is impatient or aggressive.

This echoes the central claim of the Relational Dignity Framework that dignity is co-produced in interaction and is never guaranteed by equipment or environment alone.

Together, the two frameworks suggest that technology-assisted transfers are best understood as instances of person-centered care delivery, in which the device is one contributing element among several, and where dignity remains fundamentally relational.

### 4.3 Positioning relative to existing literature

Current literature on transfer technologies primarily focuses on biomechanical criteria and reducing work-related musculoskeletal disorders (WMSDs) among staff (Bernal et al., 2015; Caponecchia et al., 2020; Hegewald et al., 2018; Martimo et al., 2008). Similarly, systematic reviews of safe patient handling interventions evaluate success primarily in terms of staff injury and implementation outcomes rather than patient experience (Wåhlin et al., 2022). While these technical advancements are important, they often overlook the ‘melancholy’ or ‘helplessness’ experienced by patients during the process. Even the limited evidence that examines patient-level outcomes generally focuses on proxy measures, such as transfer time or single-item comfort ratings (Alamgir et al., 2009a, 2009b), rather than providing an in-depth account of the patient’s experience. This study shifts the focus from professional biomechanical safety to the patient’s experience. It highlights that ‘safety’ for patients is as much about trust in the operator as it is about strap integrity or battery life. This conclusion is consistent with, and extends, qualitative dignity research in long-term care more broadly (Fuseini et al., 2023; Liu et al., 2025; Šaňáková & Čáp, 2019). This aligns with the wider literature on assistive technology, which has identified a lack of human interaction as a significant barrier to the sustained adoption of technology among older adults, alongside concerns related to privacy, usability, and stigma (Malden et al., 2025).

### 4.4 Implications for Evaluating Comfort, Dignity, and Safety

These concepts require evaluation using frameworks that incorporate the patient’s perspective. Non-verbal metrics: Professionals described reading facial expressions and body language during transfers (Section 3.2.1), which suggests that evaluation tools should not rely solely on verbal self-report.

Real-time feedback is essential for accurately capturing these cues. Evaluations should not be static; they must include “rolling consent” and real-time pain assessments (e.g. 10/10 scales) to account for daily fluctuations in patient strength or medical stability.

Stakeholder triangulation: Evaluations must involve a ‘team approach’, integrating the professional’s clinical assessment with the patient’s subjective feeling of ‘reassurance’, in line with person-centred outcome indicators that combine clinical and experiential data (McCormack & McCance, 2017).

### 4.5 Implications for Clinical Practice and Implementation

Implementing transfer technologies involves more than just installing devices. The findings suggest that comfort, dignity and safety are co-constructed through the interaction between technology, healthcare professionals and care recipients. This highlights the importance of person-centered implementation approaches.

#### Training and competency development

Participants emphasized the need for standardized, ‘just-in-time’ training that goes beyond the technical operation of transfer equipment. Educational initiatives should integrate biomechanical skills and relational competencies, such as communication, reassurance, protection of privacy, and recognition of non-verbal indicators of distress.

#### Communication during transfers

It was found that continuous communication was a key factor in ensuring comfort, dignity and safety. Healthcare professionals should provide step-by-step explanations, seek ongoing assent and consent, and use verbal and non-verbal check-ins throughout the process. Such practices may help to reduce anxiety, provide reassurance and give patients a greater sense of control.

#### Shared decision-making and patient involvement

Participants consistently emphasized the importance of receiving information, being listened to and being involved in decisions related to transfers. Wherever possible, patients and residents should be actively involved in choosing transfer methods, communicating their preferences, and identifying strategies to improve their comfort and dignity. This ensures that a patient’s unique physical, cultural and/or personal requirements are always taken into consideration.

#### Person-centered implementation

The findings suggest that the success of transfer technologies cannot be evaluated based on technical performance alone. Effective implementation requires attention to relational continuity, familiarity between staff and residents, adequate staffing levels, customized equipment, and organizational processes that support personalized care. Therefore, technology should be implemented as part of a broader person-centered care strategy, rather than as a stand-alone technical solution.

## 5. Strengths and Limitations of the Study

### 5.1 Strengths

This study has several methodological and theoretical strengths that advance the current understanding of technology-assisted transfers in long-term and complex continuing care. Firstly, it benefits from a robust dual-stakeholder and multi-site design that integrates the lived experiences of care recipients with the frontline perspectives of a multidisciplinary care team, including registered nurses, personal support workers, and physiotherapists, across multiple facilities. This provides a comprehensive understanding of transfer practices. Secondly, this study demonstrates analytical rigour by applying reflexive thematic analysis (Braun & Clarke) and adhering to COREQ-guided reporting. Reflexive journaling, analytical discussions and maintaining an audit trail supported transparency, reflexivity and methodological coherence throughout the research process. Thirdly, in a research domain that has historically centred on caregiver biomechanics and injury prevention, this study gives sustained primacy to the subjective voices of older adults. It illuminates how mechanical transfers directly affect their dignity, autonomy, and emotional security. Finally, the rich contextualized qualitative data provided an in-depth account of the transfer experience, informed a relational interpretation of technology-assisted transfers, highlighting how technologies, interpersonal care practices, and organizational conditions interact to shape comfort, dignity, and safety. This demonstrates how mechanical or technical tools, human relational practices, and organizational constraints intersect in daily care delivery.

### 5.2 Limitations

Despite these strengths, a few methodological limitations should be acknowledged. Firstly, the use of convenience sampling within healthcare facilities located in a single urban region (Ottawa, Ontario) may limit the direct transferability of the findings to rural settings, home care environments or different institutional policy contexts. In addition, as sampling prioritized availability rather than deliberate variation in characteristics such as mobility, diagnosis, or functional status, the range of profiles captured may not be fully representative. Secondly, individuals with cognitive impairment were not included, as the consent process and interview format were not adapted for this population; their perspectives therefore remain to be explored with adapted methods. Thirdly, although the study was conducted in a bilingual region, only English-speaking participants took part in the study, which may have resulted in an underrepresentation of non-English-speaking participants from diverse ethnocultural backgrounds. Additionally, while nurses and personal support workers were heavily represented, physiotherapists, who are responsible for initial transfer assessments, were underrepresented in focus groups. Finally, participants who agreed to take part may have been more attentive to transfer-related issues than the broader population of residents and staff, which may have shaped the perspectives shared. Formal member checking was not undertaken, so participants did not have the opportunity to comment on the interpretation of their accounts.

## 6. Implications for Future Research

Based on the empirical findings, the Person-Centered Practice and Relational Dignity frameworks and the methodological boundaries of this study, several coherent research priorities emerge. Firstly, future studies should design and evaluate over time “just-in-time” educational interventions that integrate relational etiquette, such as maintaining eye contact, providing step-by-step narration and decoding non-verbal cues of distress, with biomechanical instruction. Secondly, comparative clinical trials are needed to evaluate the effectiveness of specialised transfer accessories, such as pressure-relieving “Comfy slings”, customised sling dimensions for individuals with limb loss or bariatric needs, and gentler fabric materials, on skin integrity, shear force reduction and perceived comfort. Finally, implementation science research should investigate how institutional policies, staffing models and shift-to-shift communication tools, such as room-door signage and visual logos, shape transfer outcomes in various care environments, including home care, rural long-term care and acute rehabilitation units. Comparative implementation studies are particularly needed to address the tension between mandatory two-person lift policies and chronic staffing shortages, which result in delayed or suboptimal care.

## 7. Conclusion

Technology-assisted transfers should be understood not merely as technical procedures but as relational care activities. Comfort, dignity, and safety emerged through the interaction of technology, healthcare professionals, and residents, suggesting that person-centered implementation may be as important as the technology itself.

## Data Availability

All data produced in the present study are available upon reasonable request to the authors.

## Acknowledgments

The authors would like to express their sincere thanks to all the patients, residents and healthcare professionals who generously shared their experiences and perspectives. Our gratitude also goes to Michaela Adams at Perley Health and Lynsey James and Andrea Jewell at Bruyère Health, as well as the clinical leadership teams at Saint-Vincent Hospital, for their invaluable support in recruiting participants and coordinating the study. We would also like to thank the healthcare organizations that participated in the study, without whose collaboration this research would not have been possible.

## Supplementary File 5. Final thematic structure with illustrative codes

This table presents the final thematic structure reported in the Results (Sections 3.2 and 3.3) and the types of content (illustrative codes) that informed each subtheme. In keeping with reflexive thematic analysis, it is an outcome record of the analysis, not a codebook that was applied to the data, and no frequencies are reported. P = patients/residents; S = healthcare professionals.

**Overarching theme: The relational nature of technology-assisted transfers, with comfort, dignity, and safety as nested dimensions (Section 3.2).**

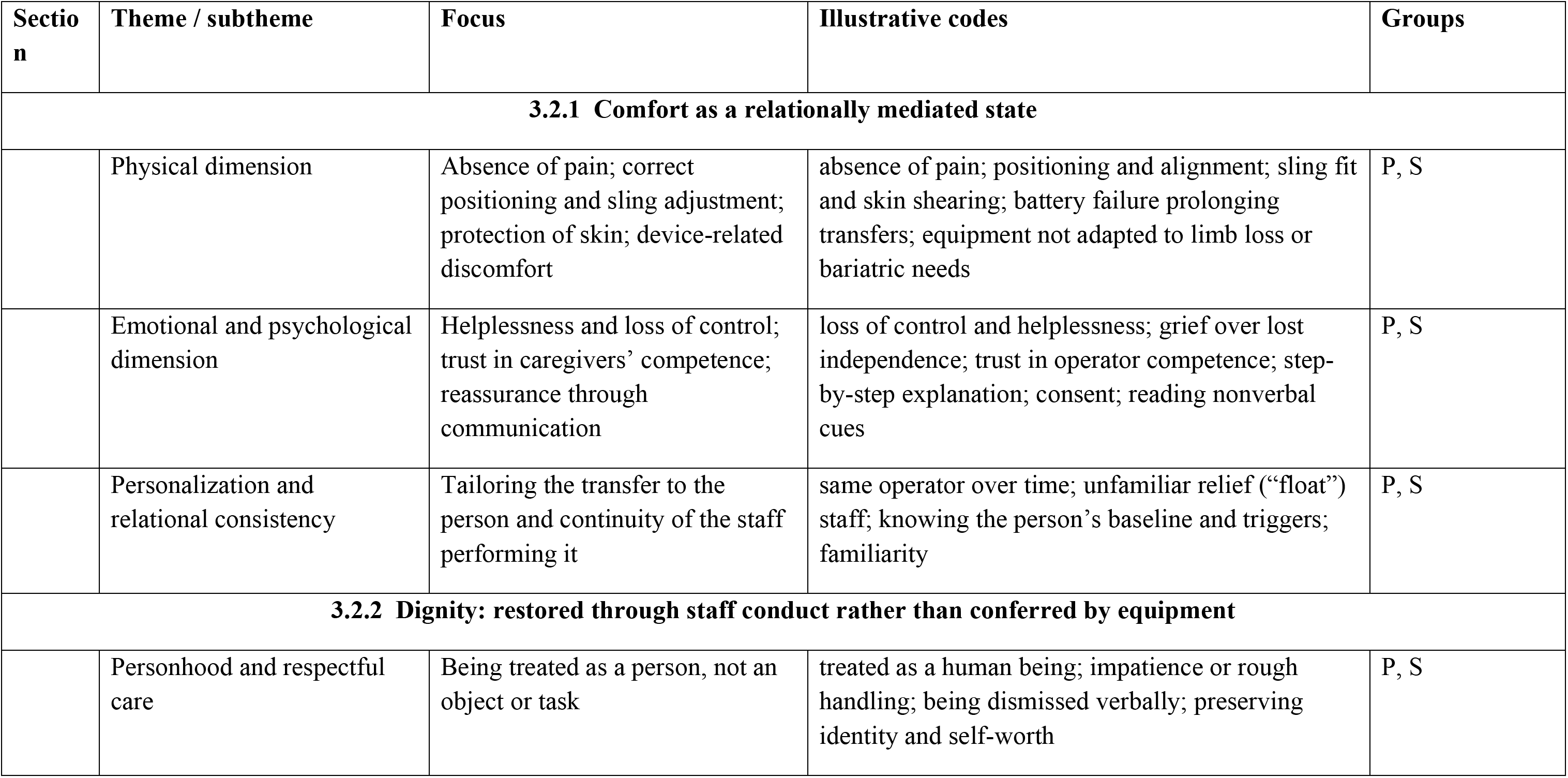
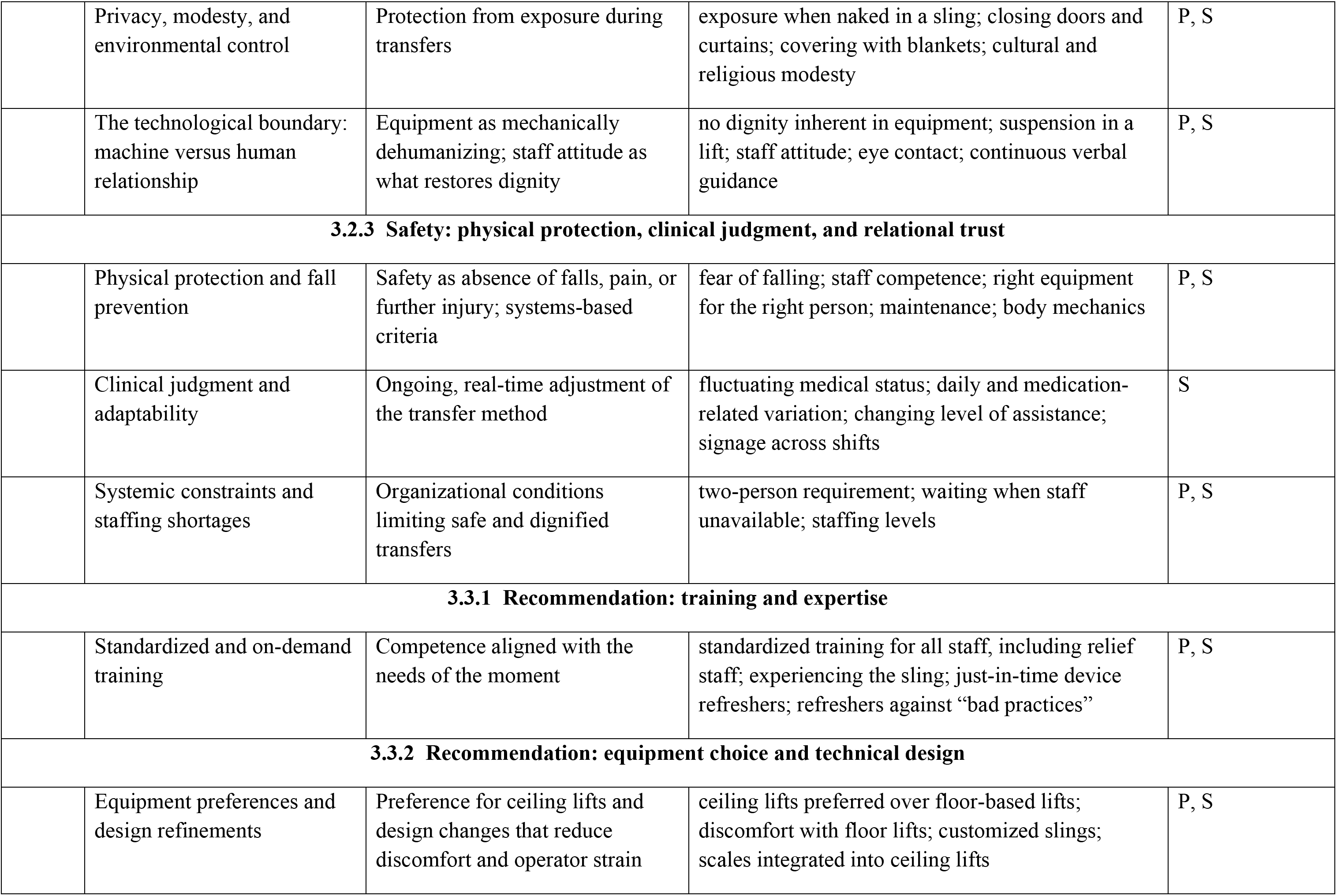
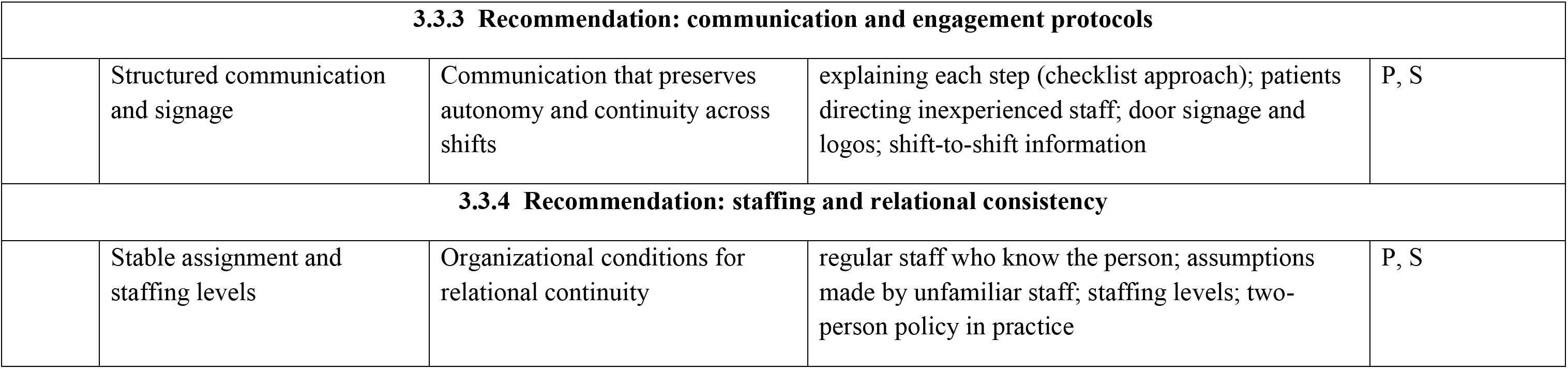

## Footnotes

1 Todam Nguepnang, N., Manirambona, L., Karunananthan, S., Toupin-April, K., Bilodeau, M., & Jutai, J. (Manuscript under review). Conceptualizing and measuring comfort, dignity, and safety in technology-assisted patient transfers: A Scoping Review.

## References

Alamgir, H., Li, O. W., Gorman, E., Fast, C., Yu, S., & Kidd, C. (2009a). Evaluation of ceiling lifts in health care settings: Patient outcome and perceptions. AAOHN Journal, 57(9), 374–380. 10.3928/08910162-20090826-06

Alamgir, H., Li, O. W., Yu, S., Gorman, E., Fast, C., & Kidd, C. (2009b). Evaluation of ceiling lifts: Transfer time, patient comfort and staff perceptions. Injury, 40(9), 987–992. 10.1016/j.injury.2008.12.002

Bernal, D., Campos-Serna, J., Tobias, A., Vargas-Prada, S., Benavides, F. G., & Serra, C. (2015). Work-related psychosocial risk factors and musculoskeletal disorders in hospital nurses and nursing aides: A systematic review and meta-analysis. International Journal of Nursing Studies, 52(2), 635–648. 10.1016/j.ijnurstu.2014.11.003

Braun, V., & Clarke, V. (2006). Using thematic analysis in psychology. Qualitative Research in Psychology, 3(2), 77–101. 10.1191/1478088706qp063oa

Braun, V., & Clarke, V. (2021). Thematic analysis: A practical guide. SAGE Publications.

Caponecchia, C., Coman, R. L., Gopaldasani, V., Mayland, E. C., & Campbell, L. (2020). Musculoskeletal disorders in aged care workers: A systematic review of contributing factors and interventions. International Journal of Nursing Studies, 110, Article 103715. 10.1016/j.ijnurstu.2020.103715

Clancy, A., Simonsen, N., Lind, J., Liveng, A., & Johannessen, A. (2021). The meaning of dignity for older adults: A meta-synthesis. Nursing Ethics, 28(6), 878–894. 10.1177/0969733020928134

Davis, K. G., & Kotowski, S. E. (2015). Prevalence of musculoskeletal disorders for nurses in hospitals, long-term care facilities, and home health care: A comprehensive review. Human Factors, 57(5), 754–792. 10.1177/0018720815581933

Fuseini, A.-G., Rawson, H., Ley, L., & Kerr, D. (2023). Patient dignity and dignified care: A qualitative description of hospitalised older adults perspectives. Journal of Clinical Nursing, 32(7–8), 1286–1302. 10.1111/jocn.16286

Hegewald, J., Berge, W., Heinrich, P., Staudte, R., Freiberg, A., Scharfe, J., Girbig, M., Nienhaus, A., & Seidler, A. (2018). Do technical aids for patient handling prevent musculoskeletal complaints in health care workers? A systematic review of intervention studies. International Journal of Environmental Research and Public Health, 15(3), Article 476. 10.3390/ijerph15030476

Heggestad, A. K. T., Høy, B., Sæteren, B., Slettebø, Å., Lillestø, B., Rehnsfeldt, A., Lindwall, L., Lohne, V., Råholm, M.-B., & Aasgaard, T. (2015). Dignity, dependence, and relational autonomy for older people living in nursing homes. International Journal for Human Caring, 19(3), 42–46. 10.20467/1091-5710.19.3.42

Jacobson, N. (2009). Dignity violation in health care. Qualitative Health Research, 19(11), 1536–1547. 10.1177/1049732309349809

Kallio, H., Pietilä, A.-M., Johnson, M., & Kangasniemi, M. (2016). Systematic methodological review: Developing a framework for a qualitative semi-structured interview guide. Journal of Advanced Nursing, 72(12), 2954–2965. 10.1111/jan.13031

Kolcaba, K. Y. (1994). A theory of holistic comfort for nursing. Journal of Advanced Nursing, 19(6), 1178–1184. 10.1111/j.1365-2648.1994.tb01202.x

Koppelaar, E., Knibbe, J. J., Miedema, H. S., & Burdorf, A. (2009). Determinants of implementation of primary preventive interventions on patient handling in healthcare: A systematic review. Occupational and Environmental Medicine, 66(6), 353–360. 10.1136/oem.2008.042481

Larsson, F., Strömbäck, U., Rysst Gustafsson, S., & Engström, Å. (2023). Perception of feeling safe perioperatively: A concept analysis. International Journal of Qualitative Studies on Health and Well-being, 18(1), Article 2216018. 10.1080/17482631.2023.2216018

Liu, Y., Wang, Y., Li, X., Ma, L., & Li, X. (2025). Dignity in nursing homes: A qualitative descriptive study of older adults’ experiences. Nursing Ethics, 32(4), 1060–1072. 10.1177/09697330241305554

Malden, S., McGill, K., Guthrie, B., Frost, H., Shenkin, S. D., Ezike, A., Bareham, B. K., Mercer, S. W., Pearce, C., Wilson, C., Underwood, I., Vines, J., Lewis, S., & O’Donnell, A. (2025). Patient and carer-related facilitators and barriers to the adoption of assistive technologies for the care of older adults: Systematic review. JMIR Aging, 8, Article e73917. 10.2196/73917

Martimo, K.-P., Verbeek, J., Karppinen, J., Furlan, A. D., Takala, E.-P., Kuijer, P. P. F. M., Jauhiainen, M., & Viikari-Juntura, E. (2008). Effect of training and lifting equipment for preventing back pain in lifting and handling: Systematic review. BMJ, 336(7641), 429–431. 10.1136/bmj.39463.418380.BE

McCormack, B., & McCance, T. (2006). Development of a framework for person-centred nursing. Journal of Advanced Nursing, 56(5), 472–479. 10.1111/j.1365-2648.2006.04042.x

McCormack, B., & McCance, T. (Eds.). (2017). Person-centred practice in nursing and health care: Theory and practice (2nd ed.). Wiley Blackwell.

Nordenfelt, L. (2003). Dignity and the care of the elderly. *Medicine*, Health Care and Philosophy, 6(2), 103–110. 10.1023/A:1024110810373

Šaňáková, Š., & Čáp, J. (2019). Dignity from the nurses’ and older patients’ perspective: A qualitative literature review. Nursing Ethics, 26(5), 1292–1309. 10.1177/0969733017747960

Sandelowski, M. (2000). Whatever happened to qualitative description? Research in Nursing & Health, 23(4), 334–340. 10.1002/1098-240X(200008)23:4-334:AID-NUR93.0.CO;2-G

Saunders, B., Sim, J., Kingstone, T., Baker, S., Waterfield, J., Bartlam, B., Burroughs, H., & Jinks, C. (2018). Saturation in qualitative research: Exploring its conceptualization and operationalization. Quality & Quantity, 52(4), 1893–1907. 10.1007/s11135-017-0574-8

Tong, A., Sainsbury, P., & Craig, J. (2007). Consolidated criteria for reporting qualitative research (COREQ): A 32-item checklist for interviews and focus groups. International Journal for Quality in Health Care, 19(6), 349–357. 10.1093/intqhc/mzm042

Wåhlin, C., Stigmar, K., & Nilsing Strid, E. (2022). A systematic review of work interventions to promote safe patient handling and movement in the healthcare sector. International Journal of Occupational Safety and Ergonomics, 28(4), 2520–2532. 10.1080/10803548.2021.2007660

World Health Organization. (2021, April 26). Falls [Fact sheet]. https://www.who.int/news-room/fact-sheets/detail/falls

World Health Organization. (2025, October 1). Ageing and health [Fact sheet]. https://www.who.int/news-room/fact-sheets/detail/ageing-and-health

